# Healthcare system barriers impacting the care of Canadians with myalgic encephalomyelitis: a scoping review

**DOI:** 10.1101/2023.09.20.23295809

**Authors:** Said Hussein, Lauren Eiriksson, Maureen MacQuarrie, Scot Merriam, Maria Dalton, Eleanor Stein, Rosie Twomey

## Abstract

**Background:** Myalgic encephalomyelitis (ME, also known as chronic fatigue syndrome or ME/CFS) is a debilitating, complex, multi-system illness. Developing a comprehensive understanding of the multiple and interconnected barriers to optimal care will help advance strategies and care models to improve quality of life for people living with ME in Canada.

**Objectives:** To: (1) identify and systematically map the available evidence; (2) investigate the design and conduct of research; (3) identify and categorize key characteristics; and (4) identify and analyze knowledge gaps related to healthcare system barriers for people living with ME in Canada.

**Methods:** The protocol was preregistered in July 2022. Peer-reviewed and grey literature was searched, and patient partners retrieved additional records. Eligible records were Canadian, included people with ME/CFS and included data or synthesis relevant to healthcare system barriers.

**Results:** In total, 1821 records were identified, 406 were reviewed in full, and 21 were included. Healthcare system barriers arose from an underlying lack of consensus and research on ME and ME care; the impact of long-standing stigma, disbelief, and sexism; inadequate or inconsistent healthcare provider education and training on ME; and the heterogeneity of care coordinated by family physicians.

**Conclusions:** People living with ME in Canada face significant barriers to care, though this has received relatively limited attention. This synthesis, which points to several areas for future research, can be used as a starting point for researchers, healthcare providers and decision-makers who are new to the area or encountering ME more frequently due to the COVID-19 pandemic.

**Funding:** This study was funded by the University of Calgary (VPR Catalyst Grant) and the Interdisciplinary Canadian Collaborative ME (ICanCME) Research Network (New Frontier ME Discovery Grant).

## Background

Myalgic encephalomyelitis (ME, also known as chronic fatigue syndrome or ME/CFS) is a debilitating, complex, multi-system illness. ME is characterized by a substantial reduction or impairment in function accompanied by profound fatigue, post-exertional malaise, unrefreshing sleep, and cognitive impairment or orthostatic intolerance [1]. The 2019 Canadian Community Health Survey (CCHS) estimates that nearly half a million people in Canada live with ME [2]. The most common onset event for ME is an acute infection, and there are known links between post-COVID-19 condition (long COVID) and ME [3]. Therefore, the current burden of ME in Canada is significantly underestimated. The pathogenesis of ME is not fully understood, and although research on the biological underpinnings is rapidly progressing, there is currently no definitive biomarker and no known cure.

Globally, ME has a long history of controversy and low credibility [4], and patients are still impacted by stigma and disbelief within the healthcare system [5]. People with ME would benefit from timely screening and diagnosis and access to expert and multidisciplinary care [1]. Yet, negative interactions with the healthcare system and significant unmet healthcare needs seem to be the norm. People with ME have described healthcare system barriers – an obstacle that prevents or restricts the use of health services by making it more difficult for individuals to access or benefit from care. The Canadian healthcare system is unique and diverse, but to our knowledge, Canadian health services research on ME is sparse. Combined with the low level of trust given to patient testimonials, this may explain why Canadian decision-makers appear to have underrecognized the legitimacy and severity of the unmet needs of people living with ME.

Developing a comprehensive understanding of the multiple and interconnected healthcare system barriers will help advance strategies and care models to improve the quality of life of people living with ME. Our approach to developing this understanding involved a synthesis of existing literature and interviews with healthcare professionals who provide care for people with ME or have lived experience of ME. This article represents one element of this methodological triangulation, and the second is reported in a companion paper [6]. A scoping review was indicated because the concept of healthcare system barriers is broad; it was unclear what information was available across peer-reviewed and grey literature, and this method of knowledge synthesis is used to identify and map the available evidence. The objectives of this scoping review were to: (1) identify and systematically map the available evidence on healthcare system barriers for Canadians with ME; (2) investigate the design and conduct of research on this topic; (3) identify and categorize key characteristics or factors related to healthcare system barriers for Canadians with ME; and (4) identify and analyze knowledge gaps.

## Methods

The Preferred Reporting Items for Systematic reviews and Meta-Analyses extension for Scoping Reviews was used to report this review [7].

### Protocol and Registration

The review protocol was prospectively registered on the Open Science Framework on July 11, 2022 (https://osf.io/fb2yk/), and no significant deviations occurred.

### Eligibility Criteria

Three concepts informed our eligibility criteria and search strategy: (1) ME (including ME/CFS or CFS, which are also used in the literature); (2) Canadian context (“Canadian” is used throughout to refer to people residing in the land now known as Canada, with an acknowledgement that much of this land is unceded territory); and (3) healthcare system barriers.

#### Inclusion Criteria

▪ The participants, population, or context explicitly included ME. Patients were diagnosed using any recognized diagnostic criteria, or patients self-reported their diagnosis. The sample could include multiple conditions, but a proportion must have ME.
▪ The participants, population, or context were Canadian. In the case of expert reviews/consensus documents that were not otherwise explicitly about Canada, the first or last author’s institutional affiliation was Canadian.
▪ The participants/population were exclusively or primarily aged ≥18 years old (or described as adults).
▪ Peer-reviewed articles and grey literature documents, including quantitative or qualitative data relevant to healthcare system barriers or expert reviews/consensus documents that explicitly discuss healthcare system barriers (where experts could be clinicians focusing on ME or representatives of ME patient associations).
▪ MSc or PhD theses, including quantitative or qualitative data relevant to healthcare system barriers (not otherwise published in the peer-reviewed literature).
▪ Published in English.

#### Exclusion Criteria

▪ The sample or context describes chronic or complex symptoms or an illness that is not ME (such as chronic fatigue, a symptom of multiple pathologies).
▪ Articles that provide inadvertent examples of, for example, stigma and stereotyping, yet do not explicitly discuss healthcare system barriers.
▪ Study or review protocols.
▪ Conference abstracts/proceedings.

Although the concept of healthcare system barriers is wide-ranging, we were interested in barriers at multiple levels, whether governance challenges and resource constraints, health system engagement, or institutional bias. These were used in our protocol as broad examples of upstream factors that result in the patient’s experience of suboptimal care.

### Information Sources and Search Strategy

The information sources and search strategy were developed with assistance from an academic librarian.

#### Peer-reviewed Literature

We searched five databases for citation and reference data across life sciences, biomedicine, behavioural and social sciences, nursing, and general academic journals. We searched MEDLINE(R), Embase, and PsycINFO via the Ovid interface, CINAHL via the EBSCO interface, and supplemented this using the Web of Science database. These formal searches were conducted on July 11, 2022. The full search strategy for peer-reviewed literature is available in the protocol (https://osf.io/ychx3/). The search strategy was conducted using a combination of two of the three key concepts: (1) ME and (2) Canadian context. The ME concept was broad enough to capture all potentially relevant records (for example, we included chronic and persistent fatigue and post-infectious or post-viral fatigue/illness as search terms) and narrowed to ME, ME/CFS or CFS during screening. The Canadian context was captured using a search filter to retrieve studies related to Canada, Canadian provinces, and the one hundred largest Canadian centres by the University of Alberta Library [8]. The third concept (healthcare system barriers) was not included in our search strategy because pilot searches confirmed that the literature was sparse enough for us to consider all Canadian literature on ME and its potential relevance to this search concept.

#### Grey Literature

We searched Canada Commons, a database that covers Canadian E-books and government and policy documents, on November 25, 2022. We also used two custom Google search engines. A custom search engine for Canadian public health information (Ontario Public Health Libraries Association) was used to search the websites of federal and provincial health departments, public health agencies, and collaborating centres [9]. A custom Google search engine designed by the University of Waterloo Library was used to search for Canadian federal, provincial, and municipal government documents and publications [10]. These custom Google searches were conducted on January 18, 2023. Based on pilot testing for relevance, two search terms were used in the final searches for both databases: “chronic fatigue syndrome” and “myalgic encephalomyelitis.” Pilot testing using other terms such as “post-viral illness,” “fatigue syndrome,” “post-viral fatigue,” and “post-exertional malaise” did not result in any unique records. In addition, our team included patient partners who were aware of (or involved in producing) grey literature relevant to this scoping review, particularly from provincial and national ME patient associations. Patient partners provided potentially eligible documents, which were included in the grey literature screening if they had not already arisen from the database searches.

### Selection of Sources of Evidence

#### Peer-reviewed Literature

Search results were de-duplicated using the Systematic Review Accelerator [11]. All categorized duplicates (ranging from extremely likely to likely duplicates) were manually checked to confirm the correct classification as duplicates. Following de-deduplication, records were imported into Rayyan, a systematic review software [12]. Titles and abstracts were independently screened for eligibility by two researchers (SH, LE), blinded to each other’s decisions during this initial process. RT reviewed all conflicts, and records considered potentially relevant were retrieved in full and assessed for eligibility. A spreadsheet was used for full-text screening. SH or LE made preliminary notes and decisions, and RT performed a subsequent screening of all full texts against the eligibility criteria. In the case of uncertainty or discrepancies between RT, SH, and LE, all authors were consulted, and a consensus was reached via review and discussion.

#### Grey Literature

For Canada Commons, due to low numbers, all records from a search of “myalgic encephalomyelitis” were exported to a spreadsheet. For the “chronic fatigue syndrome” search, the first 200 records (ordered by relevance) were exported after pilot screening indicated that the first ∼50 records were potentially relevant. Due to low numbers, all records resulting from custom Google searches were exported to a spreadsheet. Following manual de-duplication, records were retrieved in full and assessed for eligibility by SH or LE. RT reviewed all decisions, and a consensus was reached through author discussions. Unique grey literature provided by patient partners was manually added to a spreadsheet and screened against the eligibility criteria by LE and RT (no discrepancies). All authors had the opportunity to review decisions and their justifications, and consensus was reached for all records.

### Data Charting Process and Data Items

Data from eligible studies were charted using standardized tables designed for this review using an iterative process based on seed articles. LE, SH or RT charted the data, and ES, MM, and SM validated this charting. Article-level data items included the author, year of publication, research design (or article description), sample size and characteristics (where applicable), and a summary of the data or discussion relevant to healthcare system barriers, with no (or minimal) interpretation.

### Synthesis of Results

Evidence from peer-reviewed and grey literature was presented in Tables, including a summary of the healthcare system barriers discussed.

### Patient Involvement Statement

Working Group 6 of the Interdisciplinary Canadian Collaborative ME (ICanCME) Research Network was a collaboration between researchers, clinicians, and people with lived experience of ME who met during 2-hour meetings held monthly via videoconference throughout 2021-2022. A series of studies were conceptualized by the Working Group beginning in May 2021 based on the group mandate and priorities. Patient partners participated as their capacity allowed and were involved in the grant and ethics applications. MM and SM initially collated relevant literature, which became the seed articles used in developing the systematic search strategy. Three authors live with ME and were involved throughout this review (including protocol development, adjudication on full-text screening, grey literature searches, and manuscript review and editing).

## Results

The selection of sources of evidence is reported in Figure 1, divided by peer-reviewed and grey literature searches. In total, 1821 records were identified, 406 were reviewed in full, and 21 were included in this scoping review (Figure I).

**Figure I.**
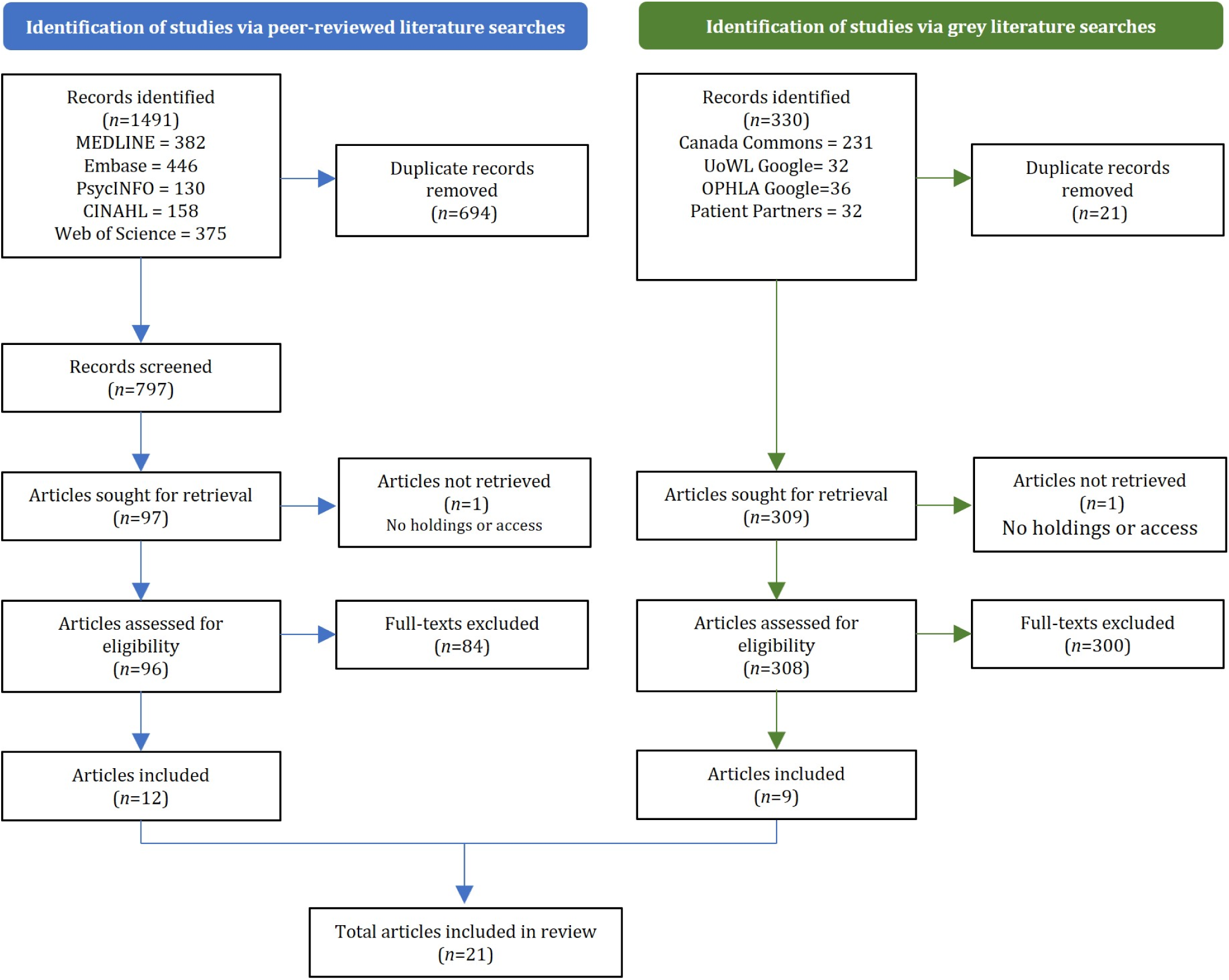
Flow diagram of the selection of sources of evidence.

### Characteristics and Results of Individual Sources of Evidence

For each article, the research design (or article description), data relevant to our research objectives, and a summary of the healthcare system barriers discussed were extracted.

Of the 12 eligible peer-reviewed articles, nine included primary or secondary data [13–21], and three were included as expert reviews/consensus documents [22–24]. Most did not explicitly focus on healthcare system barriers, and in some cases, relevance to healthcare system barriers was limited or indirect (for example, [18–20]; Table 1). One paper reported on data collected in the past decade [13]. Five peer-reviewed articles reported analyses of the data collected in the CCHS, a cross-sectional survey designed to provide population-level health information ([13–16,18]). As part of an interview, CCHS respondents were asked about specific long-term health conditions, defined as conditions that have already lasted 6 months or those expected to last 6 months, and diagnosed by a health professional. A positive case for CFS was recorded with a “Yes” in response to the question, “Do you have chronic fatigue syndrome?” Secondary analyses used CCHS cycles between 2001-2014.

Of the nine eligible grey literature articles, three reports were included as expert reviews/consensus documents [25–27]. A recent report involving a partnership between a patient association and a health centre contained a rich synthesis of the current situation for people with ME in BC alongside preliminary data [5]. Two articles were developed by an expert panel established by the Ontario Minister of Health and Long-Term Care [26,27], and the final report included two relevant appendices containing quantitative and qualitative data [26] (summarized separately in Table II). Two newsletters from patient associations presented data, one included for the analysis of CCHS data from 2014 [28] and one based on a 1992 survey of physicians in BC [29]. Two reports were Compendiums of a more extensive process guiding an Ontario Centre of Excellence Business Case involving surveys of people with ME [30] and healthcare professionals [31]. One report focused on the care and support needs of people with ME in Quebec [32].

**Table I.** Results of individual sources of evidence: Peer-reviewed literature^¥^.

| Author | Year | Title | Study design/description | Primary/ secondary data<sup>¥¥</sup> | Summary relevant to healthcare system barriers for people living with ME |
| --- | --- | --- | --- | --- | --- |
| Hu & Baines [22] | 2018 | Recent insights into 3 underrecognized conditions: myalgic encephalomyelitis-chronic fatigue syndrome, fibromyalgia, and environmental sensitivities-multiple chemical sensitivity | Commentary for Canadian family physicians | N/A | * No consistent physical or laboratory findings.<br>* Lack of proven treatments.<br>* Lack of clinical practice guidelines.<br>* Psychological symptoms mistaken for the causes rather than the effect of the disease.<br>* Uncertainty about management strategies deter clinicians and scientists from clinical care and research.<br>* Research funding "meagre or non-existent" (from 2012-2015, CIHR funded 2 grants for ME).<br>* Social stigmatization. |
| Park & Gilmour [13] | 2017 | Medically unexplained physical symptoms (MUPS) among adults in Canada: comorbidity, health care use and employment | Analysis of 2014 CCHS & 2012 CCHS-MH data in adults aged ≥25.<br>For the purpose of this study, CFS, FM, and MCS were categorized as MUPS. | *2014 CCHS data:*<br>* 1.6% [95%CI=1.4-1.8] reported CFS.<br>* CFS more common in some groups: women vs. men; without vs. with postsecondary graduation; lowest vs. higher household incomes; widowed/separated/divorced vs. married/never married; white race vs. non-white race.<br>* Comorbidities: 30% and 15% of pwCFS also had FM or MCS, respectively. 64.7% have 3 or more other chronic symptoms.<br>* Aggregate data for CFS, FM and MCS: Compared to a reference group (without CFS, FM, MCS), the aggregate group reported higher use of healthcare services (consultations with family doctors, specialists, mental health services, and other healthcare providers), unmet needs (needing but not receiving healthcare in the past 12 months), more unemployment or being permanently unable to work, more activity restrictions, including at work and at home.<br>*2012 CCHS-MH data*<br>* 35% [95%CI=27-43] of pwCFS had a mental health condition. | * In the absence of explanation for symptoms, patients may be referred to practitioners in various domains.<br>* Unmet needs despite high rates of consultation with healthcare professionals may reflect difficulties obtaining a diagnosis and treatment. |
| Bested & Marshall [23] | 2015 | Review of myalgic encephalomyelitis/chronic fatigue syndrome: an evidence-based approach to diagnosis and management by clinicians | Expert review from the viewpoint of the treating clinician, including a case study. | N/A | * No clinically available diagnostic blood test or investigation.<br>* Sparse research funding.<br>* Inconsistent ME/CFS definitions/criteria means difficulty in carrying out definitive studies that would lead to new understanding of pathophysiology, diagnostic tests, and treatments.<br>* As a result, clinicians have been skeptical that ME/CFS was legitimate, and patients have been maligned, told they did not have a real physical illness.<br>* There is a huge economic cost for the individual (income losses due to inability to work, and medical costs), their family, and society.<br>* Difficulties diagnosing ME/CFS are due to insufficient training of specialists and family doctors (most medical schools do not teach about ME/CFS in a formal systematic fashion and textbooks are not up to date in this area of medicine), fatigue being a symptom that comprises 25% of primary care physician appointments (it is imprecise and difficult to operationalize), and multiple clinical and research definitions.<br>* Studies erroneously concluded that pwCFS improve with graded exercise therapy when it can be contraindicated and harmful.<br>* Historically, cognitive behavioral therapy was inappropriately touted as a cure for ME/CFS if patients changed their "belief system".<br>* Patients often try costly alternative and complementary approaches in hopes of a cure. |
| Rusu et al. [15] | 2015 | Chronic fatigue syndrome and fibromyalgia in Canada: prevalence and associations with six health status indicators | Analysis of 2010 CCHS data in people aged ≥12. | * 1.4% [95%CI=1.3-1.6] reported CFS;<br>* CFS more common women vs. men, adults >40 vs. those aged 12-39, lowest vs. higher household incomes;<br>* Comorbidities: 23% pwCFS also had FM;<br>* Aggregate data for CFS and FM: Compared to a reference group (without these conditions), this aggregate group were more likely to report poor health status. | * Comorbidity is a central issue in the population and the cumulative effects of coexisting chronic conditions may substantially affect health status outcomes.<br>* Researchers and clinicians can anticipate substantial complexity in their studies and clinical care.<br>* Whether socioeconomic status and lifestyle risk factors are a determinant or consequence of CFS cannot be ascertained due to the cross-sectional nature of the survey. The direction of such relationships is unclear. |
| Boyd [24] | 2012 | Are some disabilities more equal than others? conceptualising fluctuating or recurring impairments within contemporary legislation and practice | Contextual discussion of the legitimacy of episodic disabilities such as ME/CFS (UK focus), drawing on ongoing discussion in Canada. | N/A | * Fluctuating or recurring impairments (episodic disabilities) such as ME/CFS are afforded questionable legitimacy, including in policy for income assistance.<br>* Although the mental or physical effects of ME/CFS are acknowledged as a set of symptoms/characteristics, the lived experience of disability as a varying phenomenon is rarely considered.<br>* Unpredictable symptoms and those not immediately visible contribute to overt suspicion, e.g., the existence and nature of ME/CFS has been long contested.<br>* A lack of physically obvious symptoms compromises recognition and identification of [those with ME/CFS as] disabled people. The unsympathetic pseudonym "yuppie flu" in the later 1980s demonstrates widespread and public skepticism.<br>* Episodic disability can have a negative impact on scheduling and coordination of care. |
| Williams et al. [16] | 2011 | Alternative health care consultations in Ontario, Canada: a geographic and socio-demographic analysis | Analysis of 2005 CCHS data in people aged ≥18 in Ontario.<br>Respondents were asked about alternative healthcare consultations (massage, acupuncture, homeopathy/naturopathy) made in the past 12months. | * People with a chronic health condition were more likely (OR=1.90) to seek alternative care than those without.<br>* People with fair/poor' health were more likely (OR=1.27) to seek alternative care than those with excellent/very good health.<br>* People with unmet needs were more likely (OR=1.72) to seek alternative care than those without.<br>* Women were more likely (OR= 2.03) to have an alternative care consultation than men<br>* In women with CFS, 42% of those with unmet needs consulted an alternative care provider vs. 24% of those without unmet needs. | * Women, people with chronic conditions, lower health status and unmet healthcare needs are more likely to seek alternative healthcare.<br>* Women with CFS are more likely to see an alternative provider if they feel their healthcare needs are not being met.<br>* Limited success in the treatment of CFS in traditional biomedical model may be one reason why alternative healthcare practices are sought out for treatment, often alongside conventional medicine. |
| Lavergne et al. [17] | 2010 | Functional impairment in chronic fatigue syndrome, fibromyalgia, and multiple chemical sensitivity. | Retrospective chart review of 128 consecutive patients with CFS, FM, or MCS at the Environmental Health Clinic at Women's College Hospital, Toronto | * N=16/72 pwCFS had paid employment and n=25/72 disability benefits.<br>* N=19/72 reported no health coverage.<br>* The main strategy pwCFS reported for dealing with health problems was rest (n=33/72).<br>* ~8-16 visits to family physicians in the past year. | * The Environmental Health Clinic is the only government-funded clinic in Ontario that is provincially mandated to assess pwCFS. It accepts referrals from family physicians and specialists, and there are long wait times for assessment. This biases patient selection towards those with greater duration and severity of illness. |
|  |  |  | (January 2005-March 2006). Canadian clinical working case definition used for diagnosis. N=72 with CFS. | * ~4-26 visits to any other physician in the past year.<br>* Participants had lower functional status relative to age and sex-matched controls, and the lowest scores were in those with combined CFS, FM and MCS. | * Unclear origin and course of CFS, and the absence of consistently abnormal physical and laboratory findings have generated difficulties for patients in securing disability support in Canada.<br>* The high number of physician visits is likely related to complexity, comorbidities, and low functional status. |
| Fuller-Thomson & Nimigon [18] | 2008 | Factors associated with depression among individuals with chronic fatigue syndrome: findings from a nationally representative survey | Analysis of 2000/1 CCHS data in adults aged ≥12. | * 36% prevalence of depression in pwCFS<br>* 31% of these depressed respondents reported that they had spoken with their family doctor about mental health issues in the preceding year.<br>* 17% had consulted a psychiatrist and 15% with a psychologist. | * Although pwCFS with depression had frequent contact with their family physician, 7/10 had not discussed mental health issues with their family physician in the preceding year.<br>* 40% of pwCFS with depression had not discussed mental health issues in the past year with any health professionals. |
| Park & Knudson [14] | 2007 | Medically unexplained physical symptoms | Analysis of 2002/3 CCHS data in adults aged ≥12.<br>For the purpose of this study, CFS, FM, and MCS were categorized as MUPS. | * CFS more common in women vs. men, those aged >45 vs. 12-44, lowest vs. higher household incomes.<br>* 43% of pwCFS needed help with instrumental activities of daily living (preparing meals, everyday housework, getting to appointments, running errands, personal finances).<br>* 15% of pwCFS needed help with personal activities of daily living (bathing, dressing, eating, taking medication, moving inside the house).<br>* 27% of pwCFS reported fair or poor mental health, 34% reported life dissatisfaction, and 36.4% reported at least one psychiatric disorder in the past 12 months.<br>* For healthcare consultations, 33% of pwCFS consulted a family doctor ≥10 times, 53% consulted a specialist, and 28% consulted an alternative practitioner in the past 12 months. | * "Medically unexplained physical symptoms" characterize conditions that cannot be identified through physical examination or medical testing, such as CFS. The lack of explanations from such assessments causes confusion and controversy for clinicians.<br>* People with "Medically unexplained physical symptoms" (such as CFS) were more likely than those without to report needing help with activities of daily living, worse physical and mental health, and more consultations with healthcare providers (which may reflect multiple referrals). |
| Fossey et al. [19] | 2004 | Sleep quality and psychological adjustment in chronic fatigue syndrome | Observational sub-study Montreal, Quebec. PwCFS recruited from physician referrals and support groups, diagnosed by physician (unknown method) and | * 58% (15/26) pwCFS fulfilled criteria for a sleep disorder such as sleep apnea/hypopnea syndrome, restless legs syndrome or periodic limb movement disorder (vs. 13% in the control group).<br>* 35/37 self-reported at least one insomnia subtype. Non-restorative sleep was common (89%). | * Prior to participating in this study, neither the pwCFS nor their physicians had been aware that they had a diagnosable sleep disorder. |
|  |  |  | confirmed by the research physician using the Fukuda definition. N=26 (analyzed). Sleep disorders assessed using polysomnography. | * Depression and neuroticism scores were in the normative range for age. |  |
| Lee et al. [20] | 2001 | Illness experience, meaning and help-seeking among Chinese immigrants in Canada with chronic fatigue and weakness. | Observational study using an adapted Explanatory Model Interview Catalogue for semi-structured interviews and numerical summaries. N=50 first-generation Chinese immigrants to Canada with prominent symptoms of medically unexplained chronic fatigue and weakness. | * 86% considered their problems to be serious, 72% reported 6-8 symptoms including fatigue, sleep disturbance, and other somatic symptoms such as headaches, muscle pains, dizziness.<br>* Simple activities of daily living were transformed into insurmountable challenges.<br>* Symptoms intensified with fatigue and developed gradually after immigration to Canada.<br>* Some felt they had never recovered from a previous severely cold winter or that they were experiencing a prolonged flu.<br>* Accounts of psychological distress emphasized impaired cognition and social functioning.<br>* Some felt unable to fulfil culturally based obligations to elderly parents and young children.<br>* Fatigue was the most frequent name participants gave to the problem (n=13) with neurasthenia second (n=10). Only two specified the common North American term CFS, and one identified the problem as "yuppie flu".<br>* Most (70%) said that they would keep others from knowing about their fatigue because it was 'a cultural thing' that Chinese people would not tell others about their "failures or disgrace". With associations to a mental illness, disclosure became an even greater concern.<br>* The advice of physicians to rest and take time off work was not appreciated since it was not a cure. Being told there was "nothing<br>wrong" was distressing rather than comforting, | * Comparative studies suggesting the prevalence of CFS is higher in white people may be due to differences in health service utilization.<br>* Little is known about CFS in specific ethnocultural groups, despite differences in the health beliefs and practices of minorities (such as the Chinese Canadians vs. the larger Canadian society). This can lead to dissatisfaction for the patient and healthcare provider.<br>* Chinese immigrants with prominent fatigue/weakness had sought help from doctors to no avail (no diagnosis or medications). They were generally dissatisfied with prior clinical care if they felt their concerns had not been heard.<br>* Participants found that traditional Chinese medicine practitioners were better able to understand the terms they most comfortably used to explain their illness, but the financial burden deterred many.<br>* Participants sought help from specialists as well as family doctors, but consultations were typically disappointing (insufficient time to listen and examine, inadequate illness explanations, short or conflicting responses to questions, treatment of isolated symptoms rather than the root cause, lack of understanding of the vocabulary of traditional Chinese medicine).<br>* Participants noted failure of healthcare professionals to understand the impact of migration, the nature of their distress, and how this adds to the frustration of an already helpless condition. |
| Clarke [21] | 2000 | The search for legitimacy and the "expertization" of the lay person: the case of chronic fatigue syndrome | Observational interview study with 60 people affiliated with Ontario-based ME/CFS support or advocacy groups.<br>Participants were either professionally or self-diagnosed with CFS (self-reported). | * 62% sought a diagnosis from >3 doctors. One reported seeing >25 doctors in a search for help. A few were diagnosed immediately because the doctor was a friend who did extra research, or because they had experience/interest in the disease.<br>* 50% were referred to a psychiatrist.<br>* 38% consulted with alternative and complementary practitioners despite costs.<br>* PwCFS developed a calculus of "good doctor" (may not be able to diagnose but believed the patient) vs. "bad doctor" (does not take symptoms seriously).<br>* 48% men and 22% women felt they had a physician who could be a regular source of information.<br>* Lay persons often become "experts" about the diseases, possible treatments, and which doctors will advocate for the patient to insurance companies and employers.<br>* Two-thirds reported that they regularly relied on their own research about symptoms, treatments, and prognosis.<br>* 78% of men and 70% of women relied upon a local support group as a source of regular information. | * The reality of CFS is contested and a substantial proportion or previous literature has a psychiatric focus.<br>* CFS has had many names and there is ambiguity in the diagnosis. A fundamental aspect of CFS is the surplus suffering experienced due to the illegitimacy of the disease and difficulties finding a doctor and a label.<br>* Most patients experience a long and complex process of getting a diagnosis.<br>* Most searches for a diagnosis involved "doctor shopping" because of personal dissatisfaction with the lack of an adequate explanation, or due to multiple referrals to organ-specific specialists resulting in multiple inconclusive tests.<br>* Differences in opinions, disputes and contradictions between doctors were observed (e.g., viral vs. psychological explanations). In these cases, the lay people were forced to decide on their own if doctors were correct or helpful.<br>* In the absence of general agreement among doctors, patients become "experts". There can be a gap between what doctors and patients know about the disease.<br>* Patients relied on and travelled to a few doctors scattered across the country because they became known to as "believers" who diagnose the disease.<br>* CFS does not readily fit in the medical model. |

CIHR=Canadian Institutes of Health Research; CCHS=Canadian Community Health Survey, a nationally representative survey; CCHS-MH=Canadian Community Health Survey-Mental Health; FM=fibromyalgia; MCS=multiple chemical sensitivities; MH=mental health; OR=odds ratio; pw=people with (such as pwCFS). \*Terminology (such as CFS and medically unexplained physical symptoms) used in the included paper/analysis has been used within this table. \*\*Primary data deemed relevant to the present research objectives.

**Table II.**
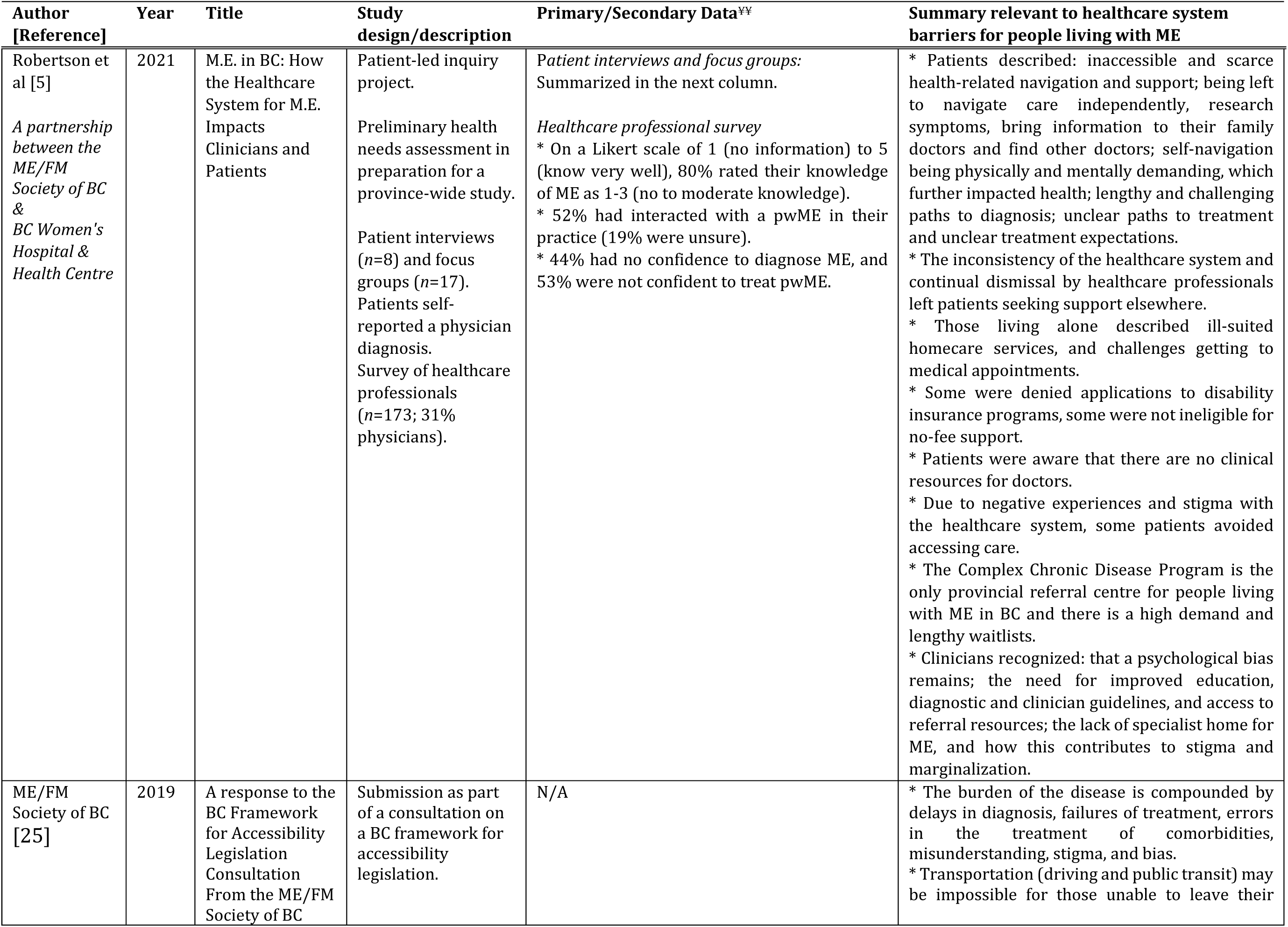
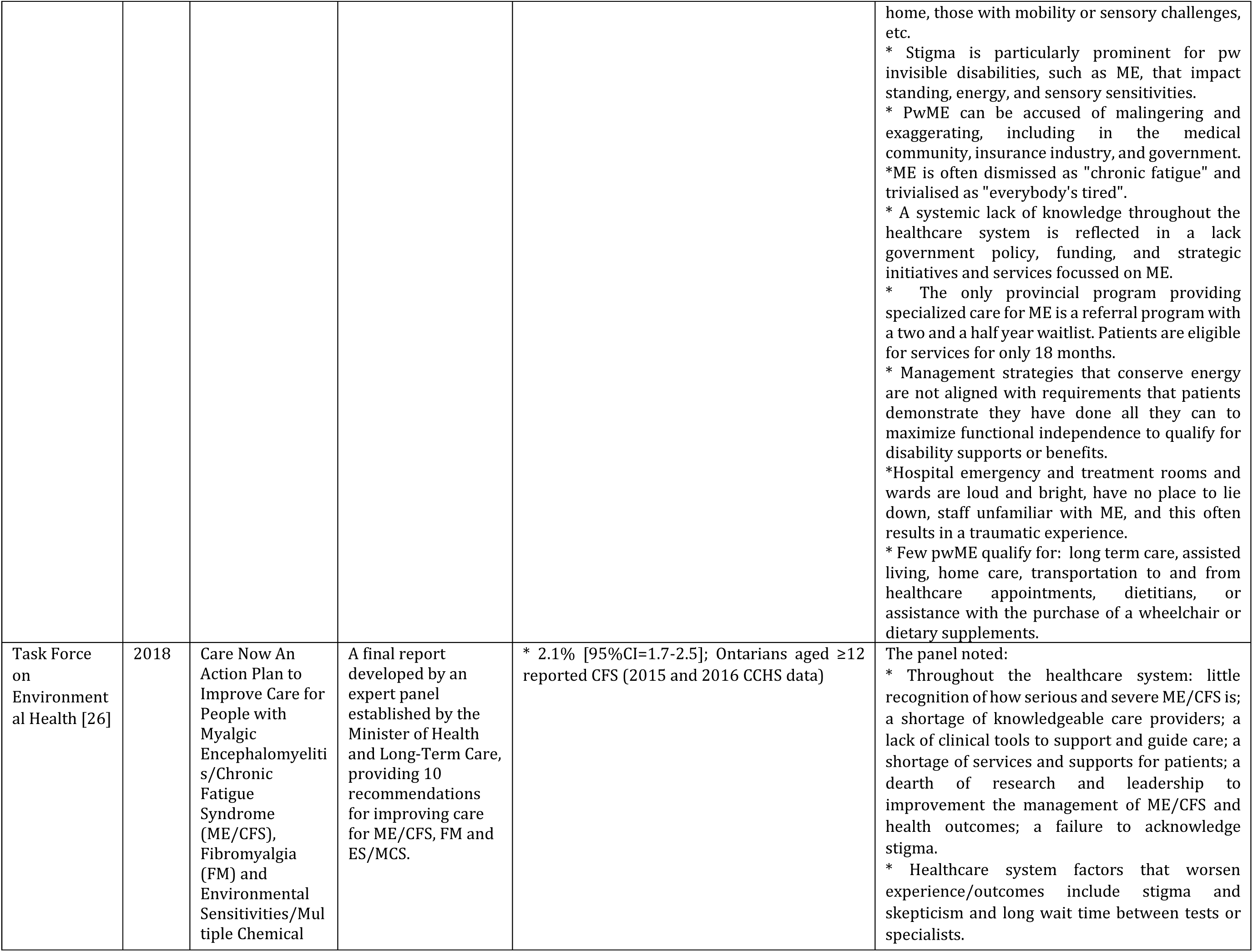
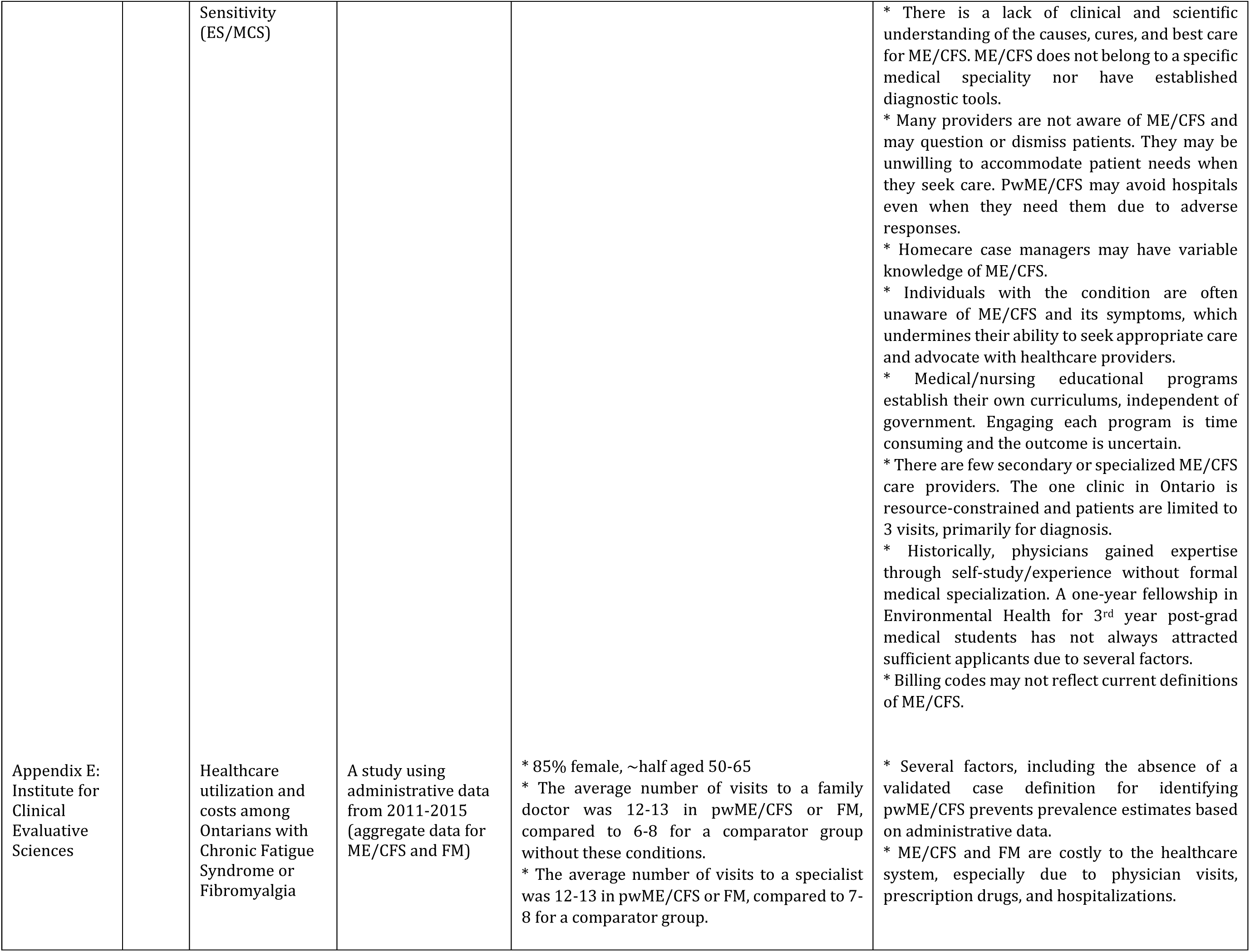
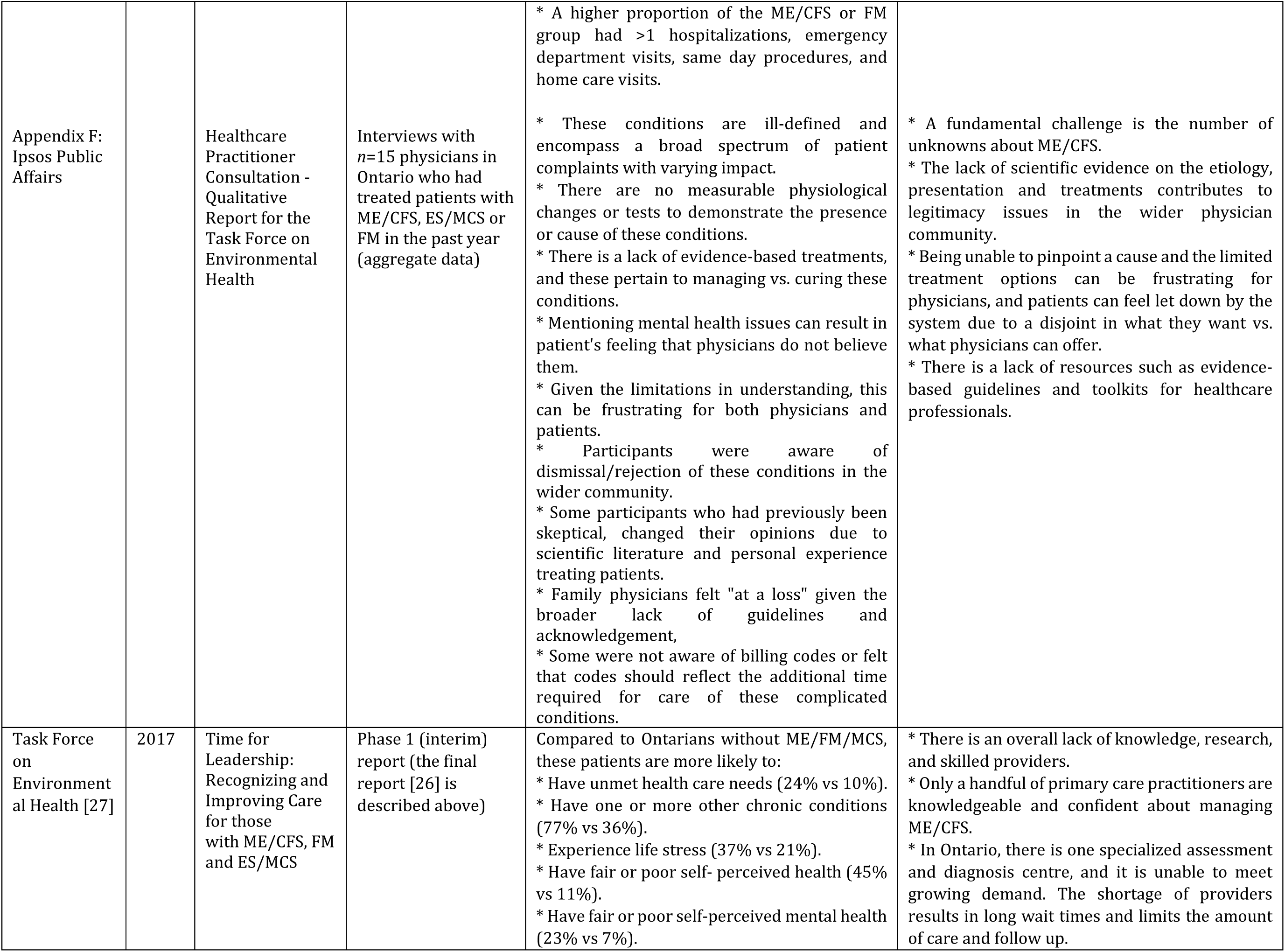
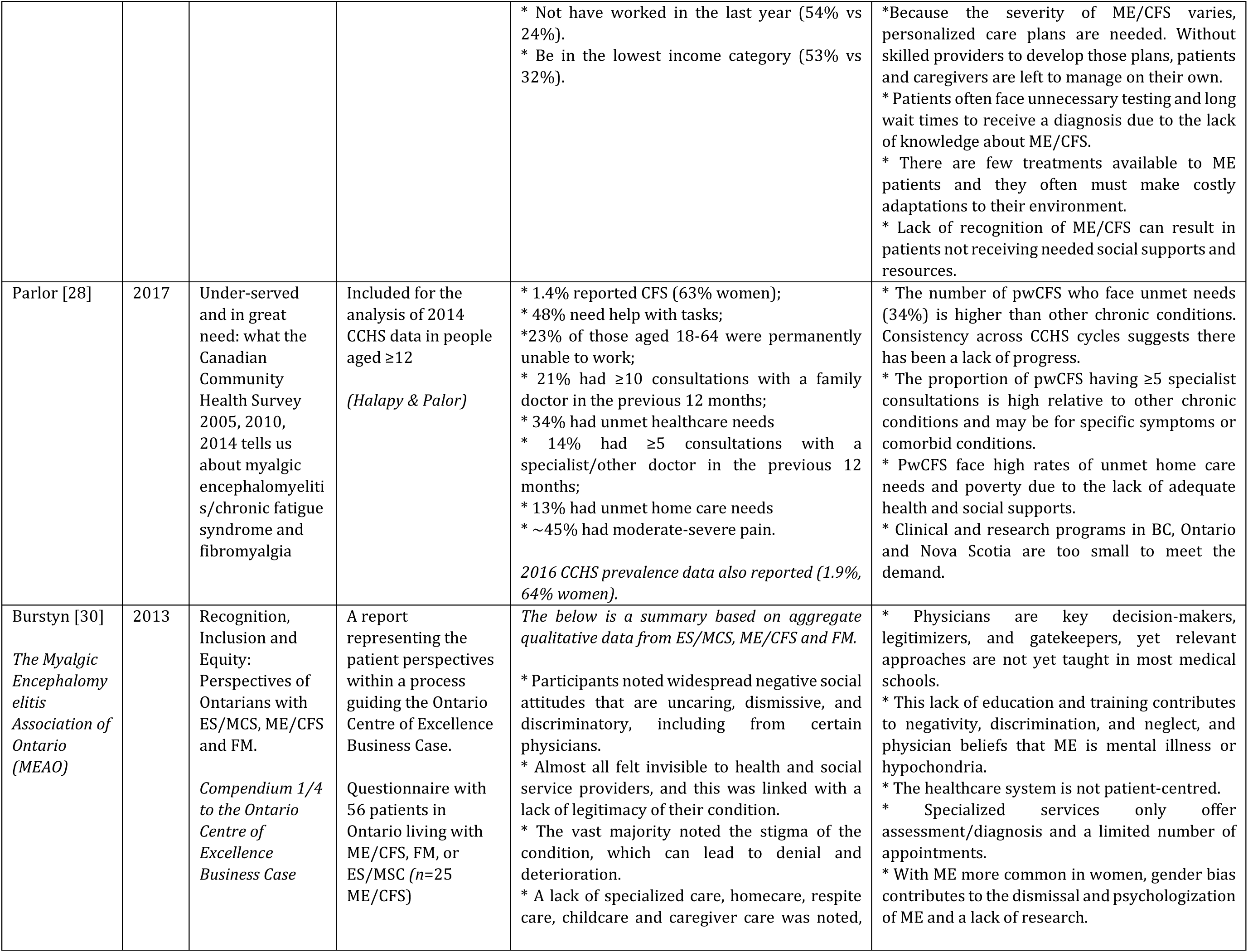
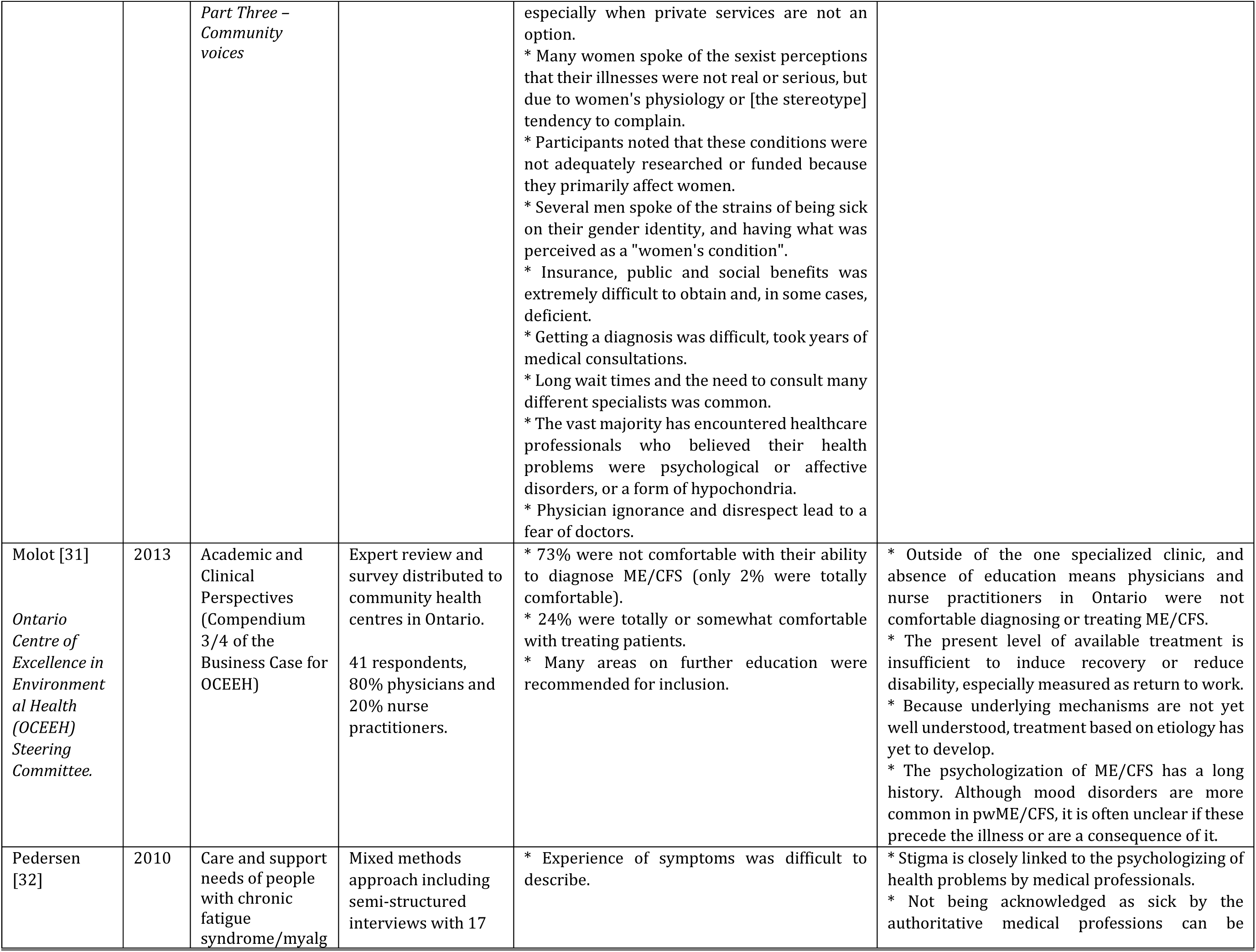
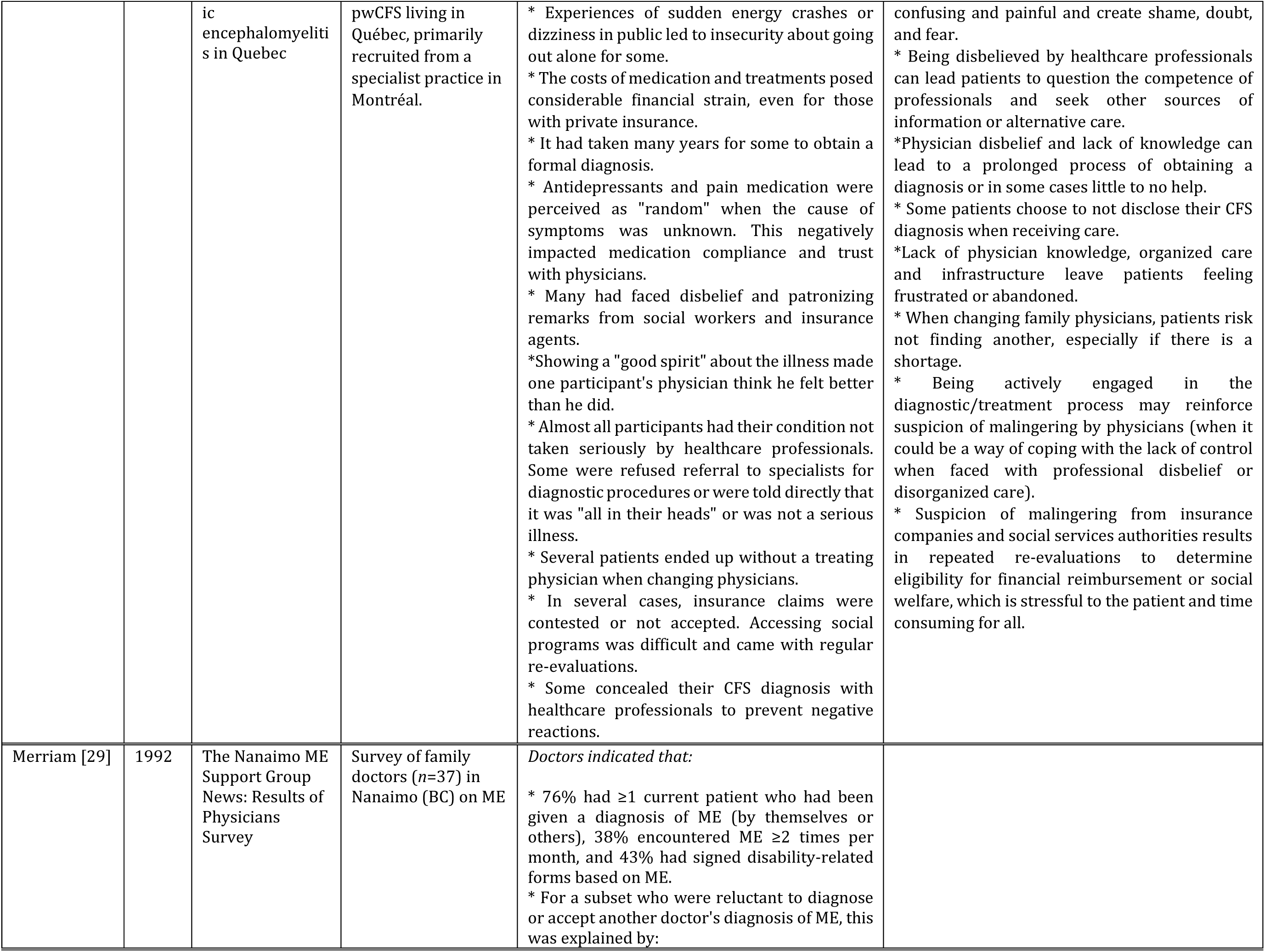
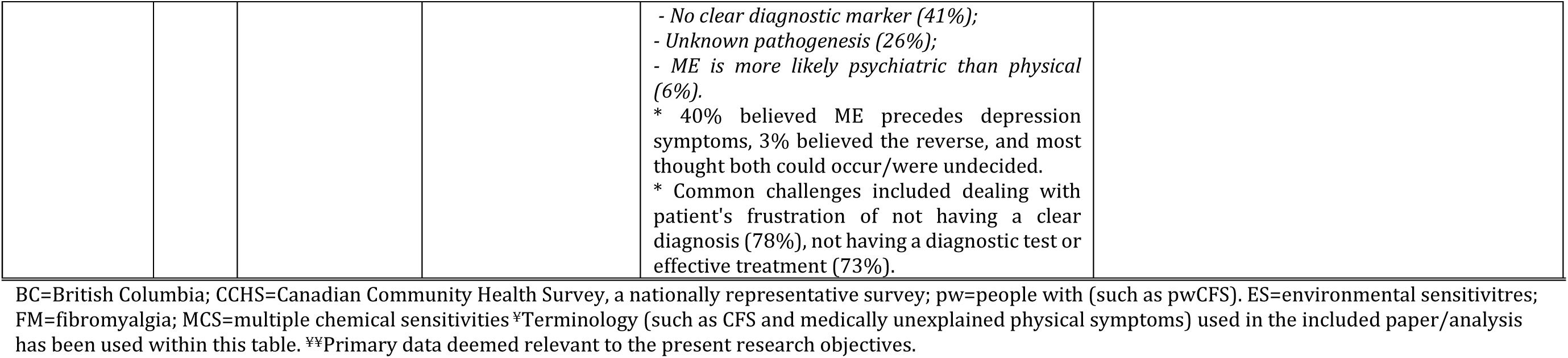
Results of individual sources of evidence: Grey literature^¥^.

Healthcare system barriers arose from an underlying lack of consensus and research on ME and ME care, the impact of long-standing stigma, disbelief, and sexism, inadequate or inconsistent healthcare provider education and training on ME, and the heterogeneity of care coordinated by family physicians. Results from peer-reviewed and grey literature are synthesized in Table I and II, respectively, and explored in more detail in the Discussion.

## Discussion

We identified the Canadian evidence relevant to healthcare system barriers for people living with ME using a preregistered scoping review process, including systematic searches of academic and grey literature and consultation with patient partners. By synthesizing the 21 eligible articles, we mapped the barriers to care reported in the literature and categorized these as outlined below (1-4). In addition, we explored the design and conduct of research on this topic. Knowledge gaps and future directions are integrated into the following sections.

### Limited available evidence on healthcare system barriers for Canadians with ME

It is telling that only one peer-reviewed article reported on data (relevant to our objectives) collected in the past decade [13]. Although overall, the literature articles contained rich and compelling information, there were methodological limitations throughout the empirical studies. With few exceptions (such as [17,26]), participant’s diagnoses were self-reported (unknown criteria), other participant characteristics were not fully described, some of the data on ME were aggregated with other conditions such as fibromyalgia [13,15,16,26,27,30], reporting standards were highly variable, and some articles involving research on human participants did not undergo ethical review [5,29]. For data on ME prevalence, healthcare utilization and unmet healthcare needs, Canada has relied on the CCHS, which has known limitations. For example, for the CCHS cycles analyzed in included articles [13–16,18,26–28], participants were asked about receiving a diagnosis of CFS by a healthcare professional. ME was not included as an option in past CCHS cycles (see [1] for more detail on naming issues related to ME and CFS). Furthermore, this data underestimates the prevalence and burden of CFS in Canada due to the difficulties, and multi-year delays patients face when seeking a diagnosis from a healthcare professional. Because the CCHS is a cross-sectional survey, it prohibits conclusions about the direction of associations, such as higher rates of depression in people with ME vs. controls [18]. Nevertheless, the assumption that depression causes or perpetuates ME appears to have gained far more traction in the healthcare system, and patient symptoms are regularly dismissed as having no biological basis [14,16,21,31,32]. There is an alternative explanation that aligns with patient testimonials - higher rates of depression are an anticipated outcome of living with a debilitating chronic illness that reduces function and limits activities.

Although there was one novel attempt to use administrative data (*Appendix E* [26]), more population health research is needed on ME. Research must be methodologically rigorous and transparently reported to generate robust and impactful evidence on ME. Given the historical issues and controversies in ME research, open science principles must be adopted, and partnerships between researchers and people with lived experience are crucial. Meaningful patient engagement goes beyond involvement as participants to involvement in governance, priority setting, developing, conducting, and/or disseminating research. The ICanCME Research Network has made progress towards this.

There are several international studies relevant to our work, and our review corroborates findings from the US, Europe, and Australia. As an example, there are published data from the US on topics such as ME healthcare utilization and economic impact [33,34], comparisons of the disparities between the burden of the disease vs. funding allocated to it [35], patient access, barriers to, and satisfaction with care [36], qualitative analyses of negative health care experiences [37]. We do not have the equivalent published data for Canada. Adding Canadian-centric evidence to the scientific record is essential for several reasons. First, this involves adhering to international standards for the conduct and reporting of research. Second, this is part of building a case for decision-makers to justify allocating more funds toward improving ME research and healthcare delivery in Canada. Third, the scientific record is where many clinicians engage in self-directed learning.

### Key characteristics or factors related to healthcare system barriers for Canadians with ME

We grouped barriers across four overlapping and interrelated categories. Patient partners noted that the situation described is an unfortunate norm they have been navigating for many years.

#### (1) An underlying lack of consensus and research on ME and ME care

Several expert reviews/consensus documents involving physicians pointed to the lack of consensus on the name, etiology, diagnostic criteria, and treatment/management of ME in the scientific literature. A lack of accepted Canadian clinical care guidelines and toolkits for healthcare professionals was also apparent. The situation has arisen from a historical lack of recognition and research funding, which has downstream impacts on available healthcare services. Even in an unprecedented scenario of a patient having early access to a knowledgeable physician who is familiar with ME, located at a sufficiently resourced clinic, and confident in assessment, diagnosis, and management, currently, there are no evidence-based treatments that cure ME or result in a return to pre-illness function.

A European Commission study concluded that ME receives insufficient research funding relative to its (high) burden [38]. The high burden of ME is assumed to, in part, stem from the lack of evidence about the etiology and treatment of the condition. The requirement to increase research activity also applies to Canada [26]. The first dedicated investment in biomedical research on ME in Canada was made in 2019, and the pathophysiology is under investigation internationally^1^. While this is underway, those involved in ME research/care have reached a consensus on several (international) clinical guidelines, and these can be shared with healthcare professionals [1,39,40]. In our experience, and based on the scientific literature, there are few independent researchers studying ME in Canada. This limits the ability of some teams to apply for funding, submit ethics applications, and publish. Alongside biomedical researchers, those with expertise in medical, educational ecosystems, anti-stigma interventions, and population health should be supported to enter the field through targeted investments.

#### (2) The impact of long-standing stigma, disbelief, and sexism

This category is not particular to Canada but arose from nearly all included resources. A historical overview was out of the scope of our review, but to summarize, ME is (and always has been) a contested illness that arouses suspicion that symptoms are exaggerated, used to avoid duties, work, or obtain insurance. On the contrary, obtaining disability insurance and other social benefits has been notoriously difficult for people with ME [5,17,25,32]. Because ME affects more women than men, the legacy of women’s symptoms being assigned to hysteria cannot be underestimated. Historically, doctors, researchers, and research participants have predominantly been male, and this is still being corrected. There are now many examples of gender bias in healthcare (the gender pain gap is one example, where women’s pain is more likely to be disbelieved, psychologized, and undertreated [41]) and gender disparity in funding of diseases predominantly impacting women [42], with ME being a prominent example. By association, this also impacts men, who are invalidated as having a predominantly female condition [30]. Stigma, disbelief, and sexism lead to healthcare visits where patient testimonials are discredited, leading to distress and withdrawal from the healthcare system to alternative care [14,16,21,32]. We point interested readers to Blease et al. [4], who describe healthcare consultations as a setting where people with ME are vulnerable to epistemic injustice.

There is evidence that people with ME are viewed differently by healthcare providers. In an analysis of an online forum, natural language processing was used to show that doctors’ attitudes are inconsistent across disease types, and ME was discussed with more negative language than all other diseases [43]. Several included articles used ambiguous terms describing symptoms as ‘medically unexplained’ [13,14,20]. Although this can be used as a broad term for symptoms for which patients seek biological care where providers have not found a biological cause, it is most often used for symptoms with primarily psychosocial causes [44]. This label can be harmful to people with complex conditions like ME because ‘unexplained’ symptoms can be used as evidence of dysfunctional illness beliefs [45]. The stress and humiliation of being disbelieved contribute to the emotional burden, which can then be incorrectly perceived as a cause rather than the consequence of the illness [32]. In addition, ME is an invisible disability that leaves some homebound or bedbound, yet for others is episodic, involving unpredictable fluctuations that occur without warning and last for uncertain periods of time. Both invisible and episodic disabilities are delegitimized, and people with ME are less likely to experience healthcare professionals who understand the support needed [24,46].

#### (3) Inadequate or inconsistent healthcare provider education and training on ME

Several articles included reports from healthcare providers noting inadequate education and training on ME [5,23,26,31], and this was reflected in patient reports [5,30,32]. In line with international studies, to our knowledge, ME is sporadically included in medical/healthcare curricula and not in sufficient depth to be comprehensive [47]. For example, in Europe, family physicians lack confidence in diagnosing or managing the care of people living with ME and desire more education [48]. With the increased awareness of post-viral illness and ME due to the COVID-19 pandemic and the involvement of occupational and physical therapists in long COVID rehabilitation, the knowledge of recently trained healthcare professionals is unknown. Although there are gaps in what we know about the current situation in Canada, the National Institute for Health and Care Excellence’s call for significant improvements in the education of healthcare professionals [40] is certainly applicable. Currently, a workforce that is experiencing unreasonable demands, high levels of stress and burnout amidst a primary care crisis [49] must identify credible sources and self-direct their learning on ME. Because this may not be possible for the majority of healthcare providers, patients have the frustrating experience of bringing research and clinical guidance to their physician [5], and this is not always received positively. Instead, a shared Canadian curriculum and series of materials to support learning could be collaboratively developed and tested. Alongside a comprehensive needs assessment for Canadian healthcare providers, models for implementing education and continuing professional development on ME are needed.

#### (4) The heterogeneity of care coordinated by family physicians

The barriers described in categories 1-3 contribute to the obstacles patients face accessing care, typically via family physicians. Depending on the knowledge, experience, and bias of the individual doctor, the experience of assessment, diagnosis, and management will vary for each patient. Not being acknowledged as sick can be confusing and painful, and multiple articles described the long and difficult paths to diagnosis that people face in Canada [5,21,26,30,32]. Even in the absence of treatments, diagnosis goes beyond validation: it is necessary for specific resources (e.g., insurance and employment services) [21]. ME is a multi-system illness that does not fit neatly under a single medical specialty and has not been widely adopted by one. Considering there are common comorbidities [13,15], and investigations are needed so other medical conditions are not missed [39], multiple referrals are often required [17,21,26,28]. Care continuity relies on an engaged and supportive family physician to help navigate the patient through the complexity of the illness and healthcare system. The family doctor can be a gatekeeper for secondary and tertiary healthcare.

The inaccessibility of care can also be an obstacle, and many patients struggle with travel, the demands of the visit, and the lack of home and virtual services. In rare specialist ME clinics (three we are aware of in Canada), patients may have a better interaction with the healthcare system, but the waitlists are long, and services are limited due to a lack of resources. Although some articles included the perspectives of healthcare professionals [26,31], the structural barriers to specializing in ME care have yet to be explored. However, some patients acknowledge the need for supportive infrastructure and appropriate reimbursement for physicians (given the complexity of CFS) [32].

### Limitations and considerations

The search may have missed relevant studies or grey literature from Québec due to a lack of resources for French translation. Because our research was focused on barriers, we did not report on the positive experiences of those who have accessed specialist care or had a supportive family doctor, but these reports (though far outnumbered by negative experiences) do exist [5,30]. Healthcare system barriers experienced by children and adolescents (and their caregivers) were not explored. Although significant overlap with the adult experience can be expected, some obstacles may be unique. In addition, participation in research is often inaccessible for people with severe ME. Although their perspectives may not be fully included herein, barriers to care are extreme. The barriers identified were drawn from samples of primarily white women (where reported) and may not be representative of the Canadian population. The experience of other genders and different equity-deserving groups living with ME is largely unknown. For example, new immigrants may experience language barriers [20], Indigenous communities lack culturally sensitive access to the healthcare system [50], and there are long-standing healthcare inequities for racialized groups, which remain to be explored in the context of ME. Systematically evaluating Canadian research inputs (funding) was out of the scope of this review but is undoubtedly an upstream factor that impacts healthcare delivery. While we provide recommendations for future health services research, recommendations made within included reports were not synthesized.

## Conclusion

People living with ME in Canada experience significant obstacles that prevent or restrict the use of health services by making it more difficult to access or benefit from care, but this has received relatively limited attention in the scientific literature. Healthcare system barriers for people with ME are numerous, interconnected, and intensify the suffering and distress caused by this chronic, complex, multi-system illness. Barriers arise from an underlying lack of consensus and research on ME and ME care, the impact of long-standing stigma, disbelief, and sexism, inadequate or inconsistent healthcare provider education and training on ME, and the heterogeneity of care coordinated by family physicians. Methodologically rigorous studies using open science and patient engagement principles are urgently needed. This synthesis, which points to several areas for future research, can be used as a starting point for researchers, healthcare providers or decision-makers who are new to the area or encountering ME more frequently due to the COVID-19 pandemic.

## Data Availability

Data produced in the study are contained in Table I and II. Online Supplementary Material can be found at: https://osf.io/ychx3/. Please contact the authors to request additional information.

https://osf.io/ychx3/

## CRediT (Contributor Roles Taxonomy)

Said Hussein: Methodology; Investigation; Writing – original draft.

Lauren Eiriksson: Investigation; Writing – review & editing.

Maureen MacQuarrie: Conceptualization; Writing – review & editing.

Scot Merriam: Conceptualization; Writing – review & editing.

Maria Dalton: Writing – review & editing.

Eleanor Stein: Conceptualization; Funding acquisition; Supervision, Project administration; Writing – review & editing.

Rosie Twomey: Conceptualization; Funding acquisition; Methodology; Investigation; Project administration; Supervision, Writing – original draft.

## Acknowledgements

From 2021-2023, several members of the Interdisciplinary Canadian Collaborative Myalgic Encephalomyelitis (ICanCME) Research Network, Working Group 6 provided input on this study. We would like to acknowledge Christiane Garcia; Izzat Jiwani; Kelli Franklin; Jane McKay; M. Sarah Selke, Farah Tabassum, and Deirdre Walsh. We would like to thank Bronte Burnette-Chiang (University of Calgary), who assisted with developing the search strategy.

## Footnotes

1 Government of Canada invests $1.4M in biomedical research to improve the quality of life of people living with myalgic encephalomyelitis: https://www.canada.ca/en/institutes-health-research/news/2019/08/government-of-canada-invests-14m-in-biomedical-research-to-improve-the-quality-of-life-of-people-living-with-myalgic-encephalomyelitis.html

